# Laboratory Epidemiology of *Salmonella* Infections and Multi-Drug Resistance Profiles in Nigeria: Barriers, Challenges and Proposed Solutions

**DOI:** 10.1101/2025.07.11.25331365

**Authors:** Amjad B.A Banibella, Nooruldeen Saad, Jafar Eyad, Ramadhani Chambuso

## Abstract

**Background:** National surveillance data on *Salmonella* antimicrobial resistance (AMR) and associated laboratory challenges are limited. There is a critical need to identify and address the barriers and challenges to strengthen the national AMR surveillance and policy guide targeted interventions for *Salmonella* infections in different geographical areas. We investigated multi-laboratory culture records to quantify AMR profiles for *Salmonella* infections, diagnostic gaps, and challenges in Nigeria.

**Methods:** Using a retrospective study, we analysed a total of 84,548 culture results from 26,630 patients across 25 public laboratories participated in the AMR surveillance report from Nigeria. *Salmonella* species and stool culture positivity rates were compared throughout the 3 years period. Stool sampling gaps were quantified and *Salmonella* species AMR for key antibiotic classes were assessed. Chi-square test and Wald risk ratios (RR) were used for statistical analysis, a p-value of **<0.05** was considered statistically significant.

**Results:** Out of 84,548 culture results, a total of 621 *Salmonella* species were isolated with *Salmonella typhi* being the most commonly reported species. Stool samples represented only 3% of all collected specimens, yet *Salmonella* species culture positivity escalated from 64% to 97% (2016 to 2017; RR 1.51, 95% CI 1.37–1.65, **p<0.001**). AMR remained entrenched: trimethoprim– sulfamethoxazole ≥90%, fluoroquinolones ≥69%, and nalidixic acid 91%; cephalosporin resistance climbed from 60% to 88%. We identified a limited stool collection compared to other samples, which impacted identification of *Salmonella* infections in an endemic area like Nigeria. The key barriers were limited laboratory data integration and lack of One Health surveillance which amplified *Salmonella* infections AMR threat.

**Conclusion:** Limited stool culture and escalating multi-drug resistance jeopardise the empirical therapy for *Salmonella* infections. Our study offers immediate, scalable interventions to strengthen One-Health *Salmonella* infections AMR control in Nigeria.

**Highlights:**

- Exceptionally high *Salmonella* positivity rate in underutilized stool samples.
- >75% resistance to core antibiotics, fluoroquinolones and cephalosporins threatens empirical *Salmonella* infections treatment.
- Limited species-level reporting on *Salmonella* cultures and low One Health *Salmonella* surveillance.
- Novel framework links microbiological gaps to targeted antimicrobial resistance (AMR) containment strategies for *Salmonella* infections.

**Significant contribution:** This work provides a rare, multi-site analysis of *Salmonella* diagnostic patterns, resistance profiles, and laboratory performance, revealing critical microbiological gaps and underutilized diagnostics potential. It contributes a novel, evidence-driven recommendations framework that integrates species-level data, AMR surveillance, and stewardship strategies, advancing precision microbiology for *Salmonella* infection control in resource-limited settings.

**Graphical abstract:** 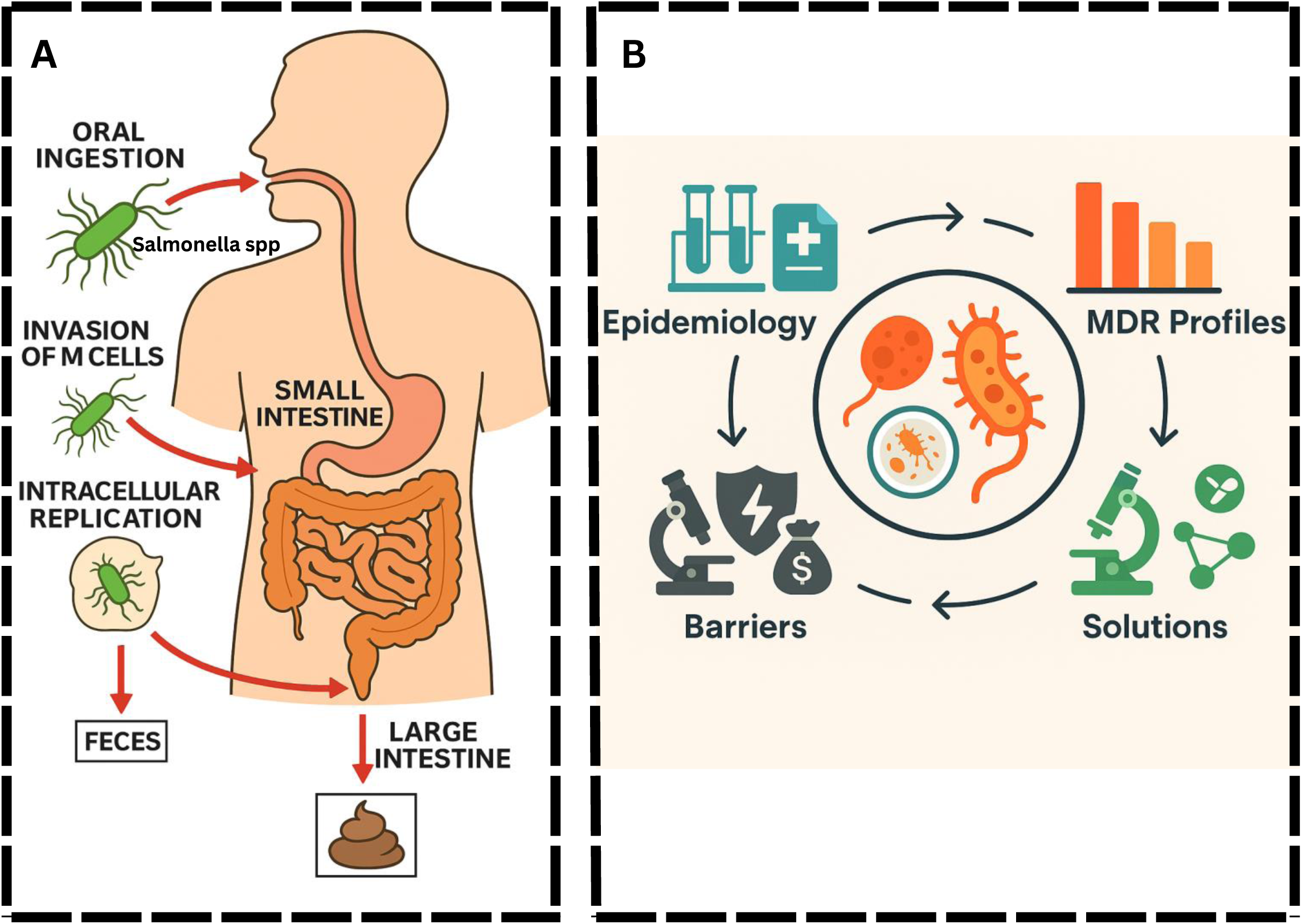

## Introduction

*Salmonella* infections remain as the leading cause of foodborne illnesses globally, with 93.8 million cases of gastroenteritis and approximately 155,000 deaths each year, of which around 80 million cases (85%) are attributed to contaminated food particularly poultry, eggs, pork, and dairy products.^1–4^ Invasive *Salmonella* infections present a particular severe form of disease causing roughly 510,000 cases and emerging as a major public health concern in 2021, with the highest disease incidence and disability-adjusted life years (DALYs) concentrated in Sub-Saharan Africa and among infants under one year of age.^5–7^ The rise of antimicrobial resistance (AMR) in *Salmonella* infections has become a critical global health concern, with a particularly severe health impact in endemic West African regions like Nigeria.^8–11^ Previous studies in *Salmonella* infections show that the whole-genome sequencing for *Salmonella* infections have revealed a high prevalence of resistance genes.^12,13^ These resistant genes were among *Salmonella* isolates from poultry, with fluoroquinolone resistance nearing 70% and widespread detection of extended-spectrum β-lactamase (ESBL) genes.^8,11,14,15^

Despite increasing reports of multidrug-resistant *Salmonella* infections in Nigeria, evidence remains limited on how the gaps in laboratory diagnostic practices can influence the accuracy and completeness of AMR detection and reporting.^16–18^ Specifically, microbiological gaps such as stool sample collection and culture for *Salmonella* infections, AMR testing protocols, and specific *Salmonella* infections policy alignment with the laboratory practices.^19,20^ There is a critical need to identify and address the barriers and challenges to strengthen the national AMR surveillance and guide targeted interventions for *Salmonella* infections in different geographical areas.^21–24^

There are two primary species of concern: *Salmonella enterica* and *Salmonella bongori*, with *S. enterica* being responsible for most human infections. *S. enterica* is further classified into over 2,500 serotypes, among which *Salmonella Typhi* is the most notable due to its role in causing typhoid fever, a severe systemic illness often found in areas with inadequate sanitation.^25–27^ *Salmonella* is a genus of rod-shaped, Gram-negative bacteria that is a major cause of foodborne illnesses. It belongs to the family *Enterobacteriaceae* and is primarily known for causing gastrointestinal infections, commonly referred to as salmonellosis.^28,29^ *Salmonella* infections drug resistance to frontline treatments such as azithromycin has reached as high as 99.4%, raising concern over treatment options for invasive infections.^30–36^ This include *Salmonella*’s ability to form biofilms, exchange resistance genes through horizontal gene transfer, and withstand environmental stresses.^37,38^

In this study, we investigated on multi-laboratory culture records to quantify AMR profiles for *Salmonella* infections, microbiological gaps including stool culture, challenges and suggested for data-driven practical solutions.

## Methodology

### Ethics statement

This study utilized publicly available, de-identified AMR surveillance data originally collected by the Nigerian Ministry of Health under the Fleming Fund Reginal Grant (Phase 1) https://aslm.org/wp-content/uploads/2023/07/AMR_REPORT_NIGERIA.pdf?x89467.

No individual-level data were accessed. Secondary data use complied with the Declaration of Helsinki and did not require additional ethical clearance.

### Study design

We conducted a retrospective study of the national AMR surveillance report of Nigeria for the years 2016 to 2018, similar to a published study.^39^ This report, published in 2022, was generated through the Mapping Antimicrobial Resistance and Antimicrobial Use Partnership (MAAP) and involved 25 sentinel laboratories across Nigeria with bacteriology testing capacity. The study adheres to the STROBE (Strengthening the Reporting of Observational Studies in Epidemiology) guidelines.^40^

### Data sources and surveillance framework

The original data were collected by the Nigerian health authorities in collaboration with MAAP partners. Surveillance activities were carried out across a national network of medical laboratories selected for their bacteriology testing capacity. A total of 25 laboratories contributed antimicrobial susceptibility testing (AST) data using WHONET software, a standardized microbiology data management tool. Data collection involved trained field teams retrieving laboratory records from both paper-based and digital systems. Where feasible, these records were linked with hospital databases to include patient demographics and clinical metadata (e.g., age, sex, specimen type).

### Inclusion criteria and data management

Only *Salmonella* isolated with valid AST results were included in the analysis. In accordance with WHO Global Antimicrobial Resistance Surveillance System (GLASS) guidelines and Clinical and Laboratory Standards Institute (CLSI) M39-A4 recommendations, only the first isolate per patient per year was retained to avoid duplicate counting. In cases where unique patient identifiers were not available, all isolates were retained and interpreted with appropriate caution. AST interpretations followed CLSI criteria, and where necessary, zone diameters or minimum inhibitory concentrations (MICs) were standardized using WHONET’s interpretive rules to ensure consistency across laboratories.

### Data extraction from the report

In this study, we manually extracted relevant *Salmonella*-specific data from the national report, including;

(i) Total number of participated laboratories
(ii) Collected specimens per year
(iii) Source of collected specimens
(iv) Specimen type
(v) Valid cultures
(vi) Positive and negative cultures.
(vii) Species-level breakdown (*Salmonella Typhi*, *S. Paratyphi*, *S. Enterica*, and *Salmonella spp.)*
(viii) AST results for antibiotics tested
(ix) Patient demographics (age and sex)

These variables were compiled into a structured Microsoft Excel dataset for a convenient data analysis. Among the *Salmonella* isolates, specific species were identified whenever possible, while others were recorded generically as *Salmonella* spp. according to how they were recorded in the original report. AMR data were available for multiple antibiotics, allowing the analysis of year-wise AMR trends and identification of the most prevalent resistant antibiotic combinations.

### AST and quality control

AST was conducted using both disk diffusion and minimum inhibitory concentration (MIC) panels, depending on the laboratory’s capacity. The primary interpretive standard applied was the CLSI criteria. Quality control (QC) strains, were widely used across labs for routine QC of AST procedures. External quality assessment (EQA) was implemented across participating laboratories via a structured proficiency testing program, coordinated with the support of MAAP and the Fleming Fund. Laboratories were encouraged to participate in periodic EQA rounds and corrective actions were documented for laboratories with poor performance. However, details on the proportion of laboratories consistently participating in EQA or specific inter-laboratory performance scores were not systematically reported.

### AMR calculation

From the original report, AMR rates in were derived from positive cultures with available AST results. AMR rates were calculated as the proportion of non-susceptible isolates (intermediate or resistant) relative to the total number of tested isolates within a single calendar year.

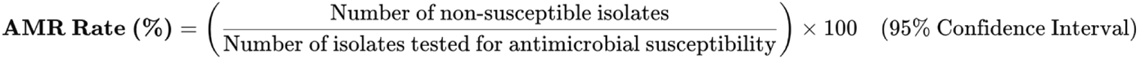

Where:

- AMR = Antimicrobial Resistance
- *Non-susceptible isolates* = Resistant + Intermediate
- *Tested isolates* = All isolates subjected to AST

### Statistical analysis

Statistical analyses were conducted using R version 4.3.2. Missing data were <1% and handled via complete-case analysis. We analysed valid culture results using descriptive statistics to compare culture positivity across demographic groups and years. For stool samples, annual *Salmonella* positivity rates were calculated with 95% Wald confidence intervals (CI), and pairwise risk ratios (RR) with 95% CI were derived using 2016 as the reference year. The Wald log method was employed to estimate RRs, and heterogeneity across years was evaluated using χ² tests. For categorical comparisons (e.g., sex, age group, year), proportions of positive versus negative cultures were assessed using Pearson’s Chi-square test, with statistical significance defined as **p < 0.05**.

## Results

### Demographics of the total cultures and the characterization of the stool culture samples

Out of 84,548 valid cultures analysed, 35.1% of female samples yielded positive results compared to 28.3% in males (**p < 0.0001**). Age-stratified analysis revealed that positivity rates increased with age, peaking at 40 % in individuals above 65 years, however, 42.5% of all culture samples had unknown age. Over time, a significant upward trend in positivity was observed from 28.5% in 2016 to 33.0% in 2017 and 30.7 in 2018 (**p < 0.0001**), suggesting improved diagnostic yield. Importantly, AST was performed for over 95% of positive cultures, ensuring robust downstream resistance profiling (Table 1).

**Table 1.** Demographics of the total cultures analayzed in the study population.

| Variable | Group | Valid cultures, n | Positive cultures, n (%) | Negative cultures, n (%) | Positive cultures with AST, n | p-value* |
| --- | --- | --- | --- | --- | --- | --- |
| Gender | Male | 37550 | 10618 (28.3) | 26932 (71.7) | 10160 | <0.0001 |
|  | Female | 46997 | 16517 (35.1) | 20828 (66.9) | 13803 |  |
| Age | < 1 year | 9689 | 2832 (29.2) | 6857 (70.8) | 2711 | <0.0001 |
|  | 1 – 17 (yrs) | 20476 | 4902 (23.9) | 15574 (76.1) | 4526 |  |
|  | 18 – 49 (yrs) | 29336 | 9323 (31.8) | 20013 (68.2) | 7629 |  |
|  | 50 – 65 (yrs) | 5177 | 1753 (33.9) | 3424 (66.1) | 1630 |  |
|  | Above 65 (yrs) | 4743 | 1898 (40.0) | 2845 (60.0) | 1806 |  |
|  | Unknown age | 15127 | 6427 (42.5) | 8700 (57.5) | 5661 |  |
| Year | 2016 | 26896 | 7678 (28.5) | 19218 (71.5) | 6532 | <0.0001 |
|  | 2017 | 32134 | 10640 (33.0) | 21494 (66.9) | 9501 |  |
|  | 2018 | 25518 | 8817 (30.7) | 16701 (65.4) | 7930 |  |

\*Pearson $\chi^2$ test comparing the proportion of positive versus negative cultures across all levels of each variable. Total number of cultures, (N = 84,548).

In order to quantify the annual *Salmonella* positivity rates in stool cultures and assess the temporal trends in *Salmonella* detection from stool samples, we analysed whether laboratory yield improved over time and if outbreak signals were emerging, or if diagnostic protocols required were strengthening. We revealing a substantial and statistically significant rise in *Salmonella* infections positivity rate from 64.2% in 2016 to over 89% in 2018 (39% higher, **p < 0.001**, **Table 2**).

**Table 2.** Characterization of stool samples for each year and the Salmonella yield.

| Year | Stool samples (n) | Salmonella-positive (n) | Positivity % (95% CI) | Pairwise Risk ratio vs 2016 (95% CI) * | p-value** |
| --- | --- | --- | --- | --- | --- |
| 2016 | 254 | 163 | 64.2% (58.1–69.8) | Reference | < 0.001 |
| 2017 | 266 | 257 | 96.6% (93.7–98.2) | 1.51 (1.37–1.65) |  |
| 2018 | 225 | 201 | 89.3% (84.6–92.7) | 1.39 (1.26–1.54) |  |
| Total | 745 | 621 | 83.4% (80.5–86.0) | n/a |  |

\*Pair-wise risk ratios (RR) were calculated with 95% CIs using Wald log-method.
\*\*Overall heterogeneity in positivity across years: $\chi^2 = 106.9$ , $P < 0.001$ (2 d.f.).

### Collected specimens distribution, culture positivity rate, and *Salmonella spp* profile

Further analyses of collected specimen patterns and laboratory performance revealed critical microbiological insights for *Salmonella* surveillance. **Fig 1a** shows that while stool specimens were consistently submitted across all years, other diagnostically relevant fluids such as bile and peritoneal aspirates remained underutilized, suggesting sampling blind spots in cases of invasive *Salmonella* infections. In **Fig 1b**, laboratory-level culture positivity ranged widely from 6.9% to 67.5%, indicating stark disparities in diagnostic performance that may compromise pathogen recovery, particularly in under-resourced sites. Despite high *Salmonella* yield, stool samples ranked only sixth among all specimen types (**Fig 1c**), reinforcing the disconnect between clinical practice and the need for stool-based diagnostics. **Fig 1d** identifies a sharp rise in *S. typhi* positive cultures in 2017 (n = 44), consistent with a potential outbreak, although species-level resolution for *S. enterica* and *S. paratyphi* remained minimal. Blood specimen submissions peaked in 2017 (**Fig 1e)**, coinciding with the rise in typhoid cases in the same year, however, stool for culture did not appear among the top five specimen types, further underscoring diagnostic underutilization. Collectively, these findings point to critical gaps in laboratory practices, specimen prioritization, and species-level identification that must be addressed to enhance *Salmonella* detection and response across endemic regions, like Nigeria.

**Fig 1.**
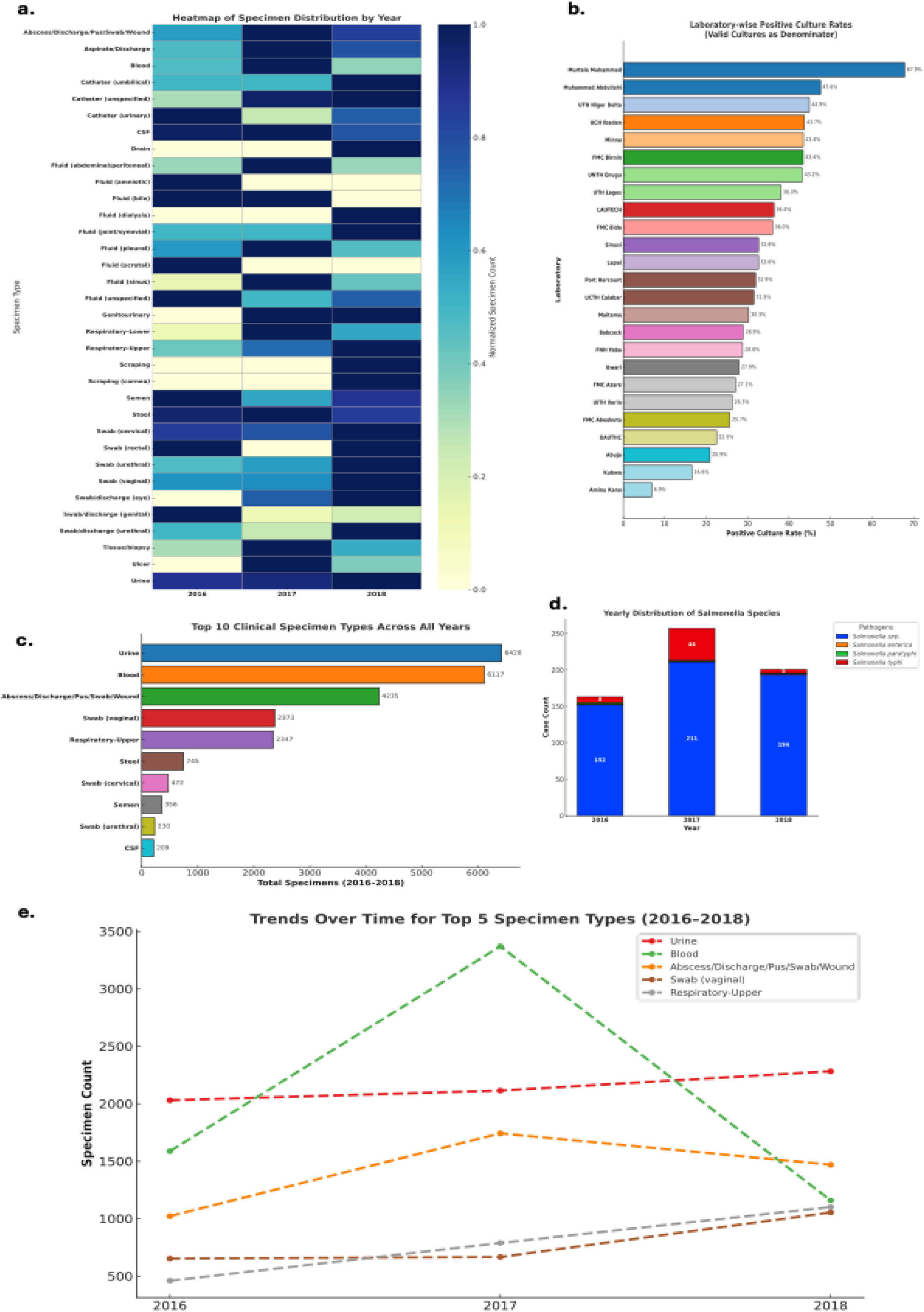
Patterns of specimen distribution, culture positivity, and *Salmonella spp.* profile. (**a**) Distribution of specimen types by year showing all specimens collected. (**b**) Laboratory-wise culture positivity showing wide variability (6.9%–67.5%. (**c**) Top ten clinical specimen types collected, showing stool samples were the sixth most collected (**d**) Yearly *Salmonella* species distribution shows a notable spike in *S. typhi* in 2017. (**e**) Trend analysis of the five most collected specimen types reveals a sharp rise and fall in blood sample collection in 2017 whereas stool remained excluded from leading collected specimen categories throughout.

### AMR surveillance in all clinical isolates and the culture for *Salmonella* Species

The AMR landscape showed a deepening crisis in the treatment of enteric infections. The overall resistance burden across the full clinical dataset shows that several key antibiotic classes such as aminopenicillins, third-generation cephalosporins, and folate pathway inhibitors exhibited resistance rates exceeding 70% (**Fig 2a**). This cross-pathogen resistance profile points to intense selection pressure and uncontained antimicrobial misuse within the system. In contrast, when we focused specifically on *Salmonella* species AMR, we uncovered a worrying temporal escalation. Between 2016 and 2018, resistance to fluoroquinolones rose sharply from 47% to 69%, while quinolone and first-generation cephalosporin resistance reached 91% and 88%, respectively, by 2018 (**Fig 2b**). Notably, even in 2016, *Salmonella* isolates already showed near-complete resistance to folate pathway inhibitors (98%) and concerning levels to third-generation cephalosporins (60%) (**Fig 2b**).

**Fig 2.**
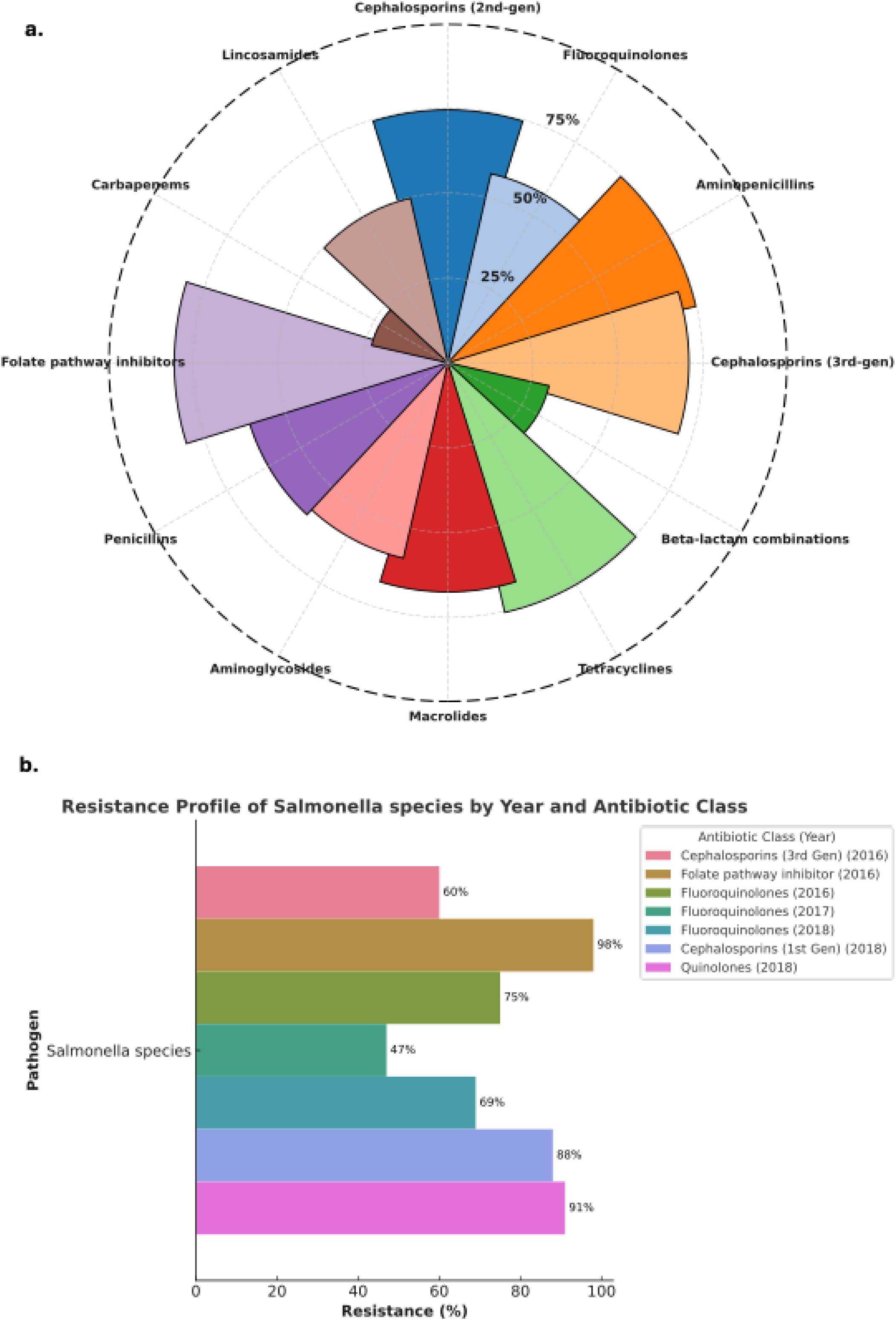
AMR surveillance in all clinical isolates and *Salmonella* Species. (**a**) Presents the overall antimicrobial resistance profile across all clinical pathogens in the dataset, revealing highest resistance rates against folate pathway inhibitors, aminopenicillins, and third-generation cephalosporins. (**b**) Shows specifically on *Salmonella* species on the marked temporal rise in resistance: fluoroquinolone resistance increased from 47% in 2017 to 69% in 2018

### Critical microbiological and surveillance gaps undermining effective *Salmonella* AMR control

To better contextualize the critical microbiological challenges surrounding *Salmonella* spp. AMR, we summarized key microbiological and surveillance gaps identified. This synthesis analysed six interlinked gaps that, taken together, impede effective detection, treatment, and containment of drug-resistant *Salmonella* strains. These gaps span between both microbiological and system-level domains, including resistance to key antibiotic classes, under-prioritization of stool sampling, and lack of integrated One Health surveillance (**Fig 3**). We highlighted what remains neglected in *Salmonella* AMR response efforts across clinical and public health settings. The observed gaps include:

(i) Fluoroquinolone (FQ) resistance (46 – 75 %) Resistance to fluoroquinolones span from 46 % to 75 %, indicating that nearly half to three-quarters of *Salmonella* isolates tested were non-susceptible to this drug class.
(ii) Invasive non-Typhoidal *Salmonella* (iNTS) in co-Infections A distinct category flags invasive NTS associated with co-infective presentations. This was recorded as a discrete surveillance element, separating iNTS through bloodstream diagnosis from the broader pool of gastrointestinal cases.
(iii) Surveillance data gaps There was observed surveillance data gap confirming incomplete or missing laboratory information, specifically, deficiencies in detailed isolate characterisation (e.g., species, serovar, and susceptibility metadata).
(iv) Trimethoprim–Sulfamethoxazole (TMP-SMX) resistance (> 90 %) Resistance to TMP-SMX surpasses the 90 % threshold, signifying that fewer than one in ten *Salmonella* isolates remained susceptible to this long-standing drug.
(v) Absence of integrated One-Health surveillance The “One-Health surveillance” was recorded as a gap, denoting that coordinated data capture across human, animal, and environmental sources was insufficiently documented for *Salmonella*.
(vi) Stool sample collection for *Salmonella* culture and AST Stool sampling was explicitly not in top 5 most collected samples for culture, confirming that, despite its relevance for enteric pathogens, stool was collected less frequently than at least five other specimen categories across the study years (**Fig 3**).

**Fig 3.**
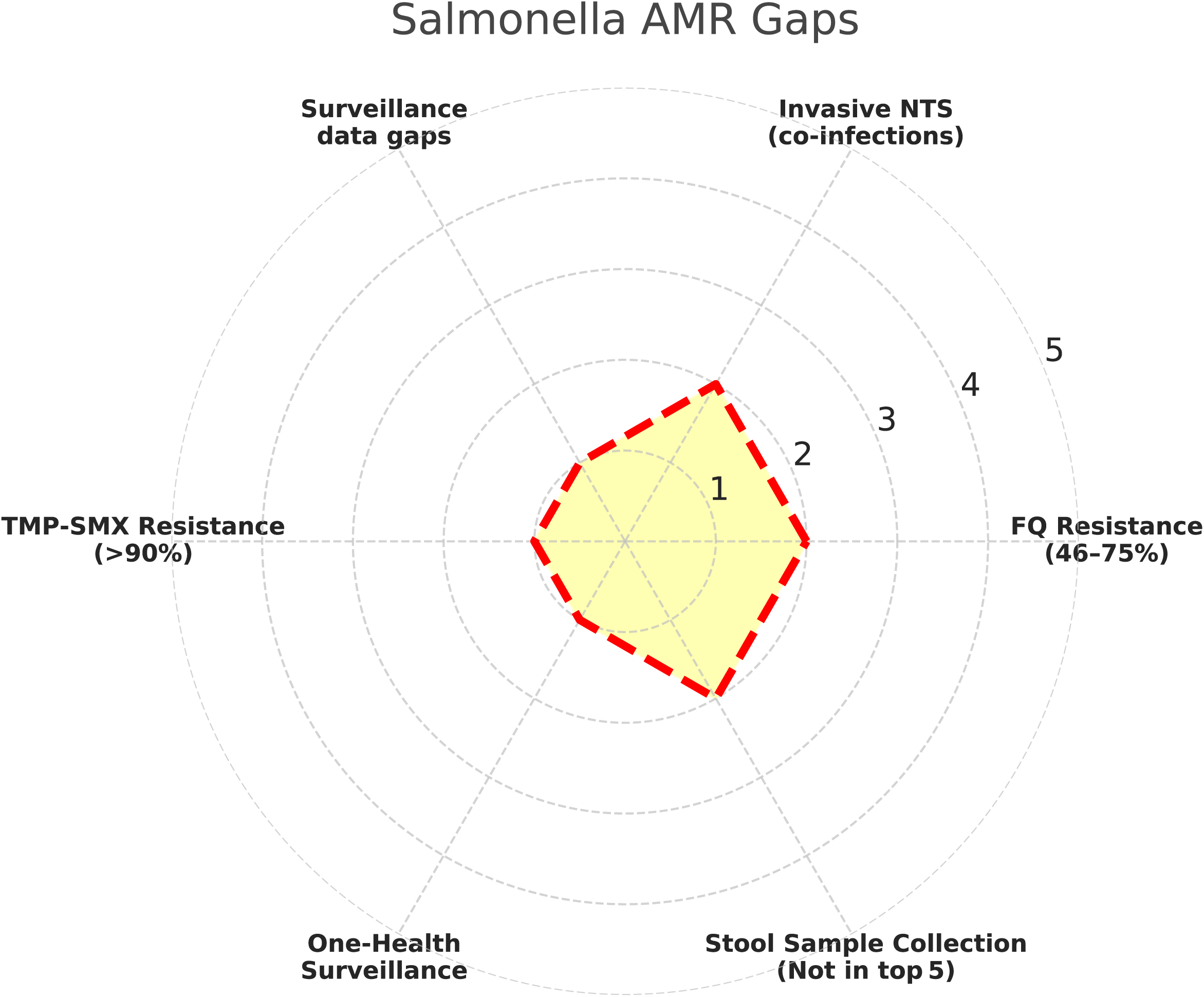
Analysis of microbiological and surveillance gaps undermining *Salmonella* AMR control. Six distinct gaps identified for *Salmonella spp*. Fluoroquinolone resistance was recorded within a range of 46–75%. Invasive non-typhoidal *Salmonella* (iNTS) associated with co-infections was identified as a separate category of concern. Surveillance data gaps were evident, particularly in species-level reporting. Trimethoprim-sulfamethoxazole (TMP-SMX) resistance exceeded 90%. Limited evidence of integrated One Health surveillance across human, animal, and environmental sectors was observed. Stool sample collection did not appear among the top five most collected specimen types.

### Microbiological recommendations for strengthening *Salmonella* AMR control

To translate the observed microbiological gaps into actionable priorities, a targeted framework of laboratory-informed recommendations was developed (**Fig 4)**. This approach systematically maps each challenge ranging from AMR and underdiagnosis to surveillance fragmentation against practical interventions rooted in microbiological best practices. These recommendations aim to improve *Salmonella spp.* detection, guide treatment decisions, and enhance cross-sectoral AMR surveillance capacity, particularly in high-burden and resource-limited settings. The recommendations are as follows:

(i) Integrate stool sample collection into AMR protocols. Routine stool sampling must be embedded into national AMR surveillance protocols, particularly for suspected *Salmonella* infections. Stool specimen is essential for identifying gastrointestinal *Salmonella* strains, yet underutilization, previously demonstrated by its absence from the top five specimen types, undermines pathogen recovery. Integration into diagnostic algorithms will improve case detection, enable species-level typing, and ensure more representative resistance profiling.
(ii) Enforce stewardship Laws to stop off-label fluoroquinolone use in poultry. Widespread off-label use of fluoroquinolones in poultry contributes significantly to the development of resistant *Salmonella* strains, which may be transmitted to humans via food or environmental routes. Enforcing antimicrobial stewardship regulations targeting veterinary sectors is crucial to reduce selection pressure and prevent further resistance amplification in zoonotic reservoirs.
(iii) Remove TMP-SMX from empiric *Salmonella* protocols and revise national guidelines. Given the AMR of >90% observed for TMP-SMX, national treatment guidelines should be updated to exclude it from empirical therapy for suspected *Salmonella* cases. This change is critical to prevent ineffective treatment, mitigate resistance escalation, and redirect clinicians toward evidence-based, susceptibility-guided *Salmonella* treatment.
(iv) Operationalize national AMR surveillance across human, veterinary, and poultry sectors. To capture the full epidemiological context of *Salmonella* transmission, surveillance must be extended beyond clinical settings to include veterinary and poultry sectors. This recommendation aligns with a One Health framework, ensuring coordinated sampling, data integration, and resistance monitoring across reservoirs that perpetuate resistant *Salmonella* strains.
(v) Develop WHONET-linked *Salmonella* module covering foodborne diseases AMR. A dedicated *Salmonella* module within the WHONET platform would enable centralized, standardized tracking of resistance trends in foodborne disease isolates. This module would allow microbiologists to monitor susceptibility shifts in real time, detect emerging resistance phenotypes, and correlate food source data with clinical outcomes.
(vi) Integrate iNTS surveillance in national sentinel disease sites. iNTS infections often associated with bloodstream infections in immunocompromised individuals require dedicated monitoring within sentinel sites. Integration of iNTS surveillance into national systems will enable early detection, facilitate targeted intervention, and support the development of diagnostics and vaccines specific to this under-recognized but clinically significant form of *Salmonella* infections (**Fig 4)**.

**Fig 4.**
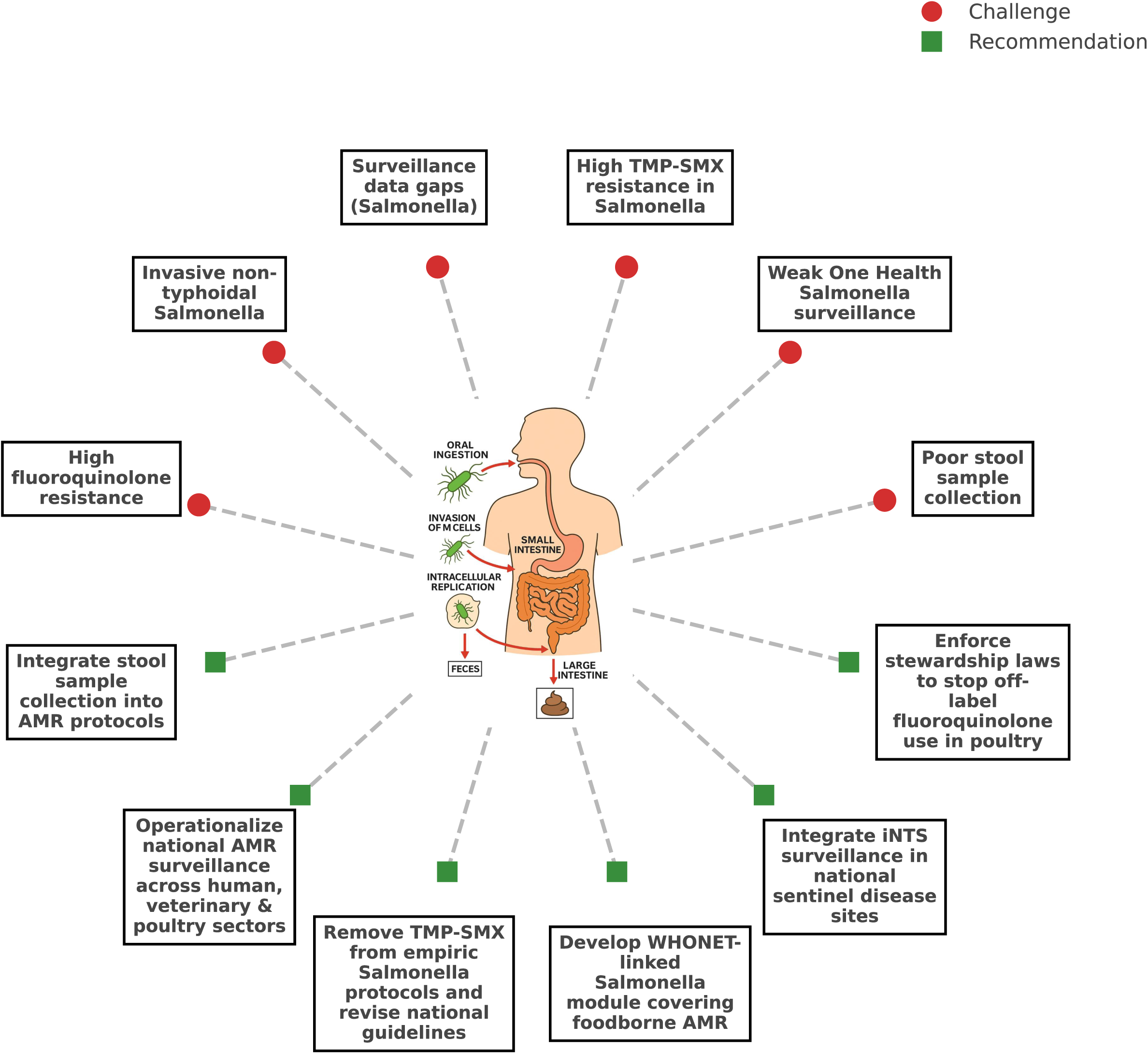
Microbiological challenges and strategic recommendations to strengthen *Salmonella* AMR control. This conceptual framework analysed form the data to illustrate potential nine microbiological and surveillance challenges surrounding *Salmonella* antimicrobial resistance (red circles), each linked to targeted, actionable recommendations (green squares). This synthesis bridges clinical microbiology, AMR policy, and pathogen ecology, offering a roadmap for strengthening diagnostic prioritization and containment strategies in endemic settings.

## Discussion

We analysed a total of 84,548 valid clinical cultures from Nigeria collected from 2016 to 2018. The specimen isolates distribution for *Salmonella* infections, detection patterns, AMR profiles, and surveillance gaps were systematically analysed across multiple diagnostic laboratories. We revealed several novel insights: despite the highest *Salmonella* culture positivity, stool samples remained significantly underutilized; species-level identification was largely absent or under reported; and fluoroquinolone resistance in *Salmonella* rose sharply, surpassing 69% by 2018. We also identified significant inter-laboratory variation in culture positivity (6.9%–67.5%) and extremely high TMP-SMX resistance (>90%), both previously underreported in regional AMR surveillance. Importantly, we developed a novel microbiological-driven gap analysis and recommendations framework linking diagnostic gaps to targeted interventions, including WHONET integration, empirical therapy revision, and One Health surveillance. This translational approach offers a scalable model for enhancing clinical microbiology for *Salmonella* control and AMR containment in low-resource, high-burden settings like Nigeria.^41–46^

These findings underscore a critical disruption in the microbiological ecosystem of *Salmonella* spp., where diagnostic underperformance and limited AMR evolution are converging. The consistent lack of species-level identification hampers the understanding of circulating *Salmonella* lineages, masking shifts in virulence and resistance phenotypes that are essential for guiding outbreak responses, vaccine targeting, and phage therapy design.^47–50^ In addition, the rise of multidrug-resistant *Salmonella* compromises the utility of both oral and parenteral treatment regimens. This may narrowing therapeutic windows and increase reliance on last-resort agents. This reflects the limited spread of mobile *Salmonella* genetic elements and clonal expansion of resistant strains, which, if unaddressed, could establish persistent reservoirs across clinical, foodborne, and environmental niches.^35,51^

Previous literature on *Salmonella* infections in Nigeria has largely focused on outbreak descriptions, bloodstream infections, or molecular typing in isolated settings, with limited integration of diagnostic workflow data, stool specimen utilization, and real-time AMR patterns.^52–59^ Our study bridges that gap by revealing the operational disconnect between high-yield diagnostics such as stool cultures and their actual underutilization in routine practice. While previous reports have documented rising fluoroquinolone resistance, our study provides robust temporal and cross-laboratory evidence of its progression, supported by context-specific resistance thresholds across antibiotic classes.^60^ Furthermore, the striking absence of species-level *Salmonella* identification aligns with known challenges in LMIC microbiology but has rarely been quantified across systems. This work adds value by providing the first structured linkage between microbiological failure points and practical, lab-driven interventions, thereby elevating the role of microbiologists from passive reporters to active architects of AMR response and surveillance reform.^61^

We exposed hidden microbiological failures in *Salmonella* infection detection and resistance containment gaps that traditional surveillance studies often overlook. By triangulating specimen practices, pathogen recovery rates, and *Salmonella*-specific resistance data, we reveal how stool under-sampling, poor species resolution, and inconsistent laboratory performance collectively distort the microbiological landscape of enteric fever caused by *Salmonella* infections.^62^ Unlike previous studies limited to single-centre or outbreak reports, this work unifies fragmented data into an actionable framework, making visible the structural and diagnostic blind spots that perpetuate *Salmonella* misdiagnosis and AMR progression.^63^ This integrative approach provides microbiologists with a blueprint for reforming diagnostic priorities and surveillance architecture in *Salmonella* endemic regions.^64,65^

We translated microbiological data into pragmatic interventions that are both context-sensitive and scalable. By aligning laboratory findings with system level diagnostics and policy gaps, our gap analysis study enables targeted improvements such as integrating stool sampling into AMR protocols, eliminating ineffective antibiotics from empirical regimens, and operationalizing One Health surveillance platforms.^66–68^ Our recommendations are practical and data oriented, derived directly from field-level diagnostic perspectives, ensuring high feasibility and translational relevance. Importantly, the new framework proposed in this study serves not only as a diagnostic audit tool but also as a decision-making scaffold for health ministries, AMR committees, and laboratory networks seeking to enhance *Salmonella* control through sustainable microbiology-led interventions for resource-limited countries.^11,69,70^

Key strengths of this study include:

(i) First to link diagnostic performance with *Salmonella* treatment gaps by establishing a unique correlation between underdiagnosis, resistance evolution, and policy inaction in endemic contexts.
(ii) Large multi-year, multi-site dataset from *Salmonella* infection endemic region. Analysis of 84,548 valid cultures across multiple diagnostic centres over three years provides robust, generalizable findings.
(iii) Microbiologically-focused gap analysis study which identified a novel framework for diagnostic and surveillance blind spots specific to *Salmonella* infections, including stool specimen prioritization, species identification, and AMR trends.
(iv) Time-trend AMR profiling which shown the escalating *Salmonella* resistance to fluoroquinolones and cephalosporins in a temporal, multi-centre context from a LMIC setting.
(v) Actionable laboratory-based recommendations focused on microbiology and directly translated findings into implementable solutions such as empirical treatment revisions, WHONET integration, and One Health coordination in real world.
(vi) New systems-level diagnostic insights which integrated specimen workflow data with microbiological outcomes specific for *Salmonella* infections, exposing variability in stool culture positivity and diagnostic capacity for each year.
(vii) Translational relevance to public health by providing microbiological evidence to inform *Salmonella* infections AMR combating strategies, health ministries, and laboratory networks seeking to enhance Salmonella infections surveillance and treatment policies in limited-resource countries.
(viii) Scalable and reproducible methodology which offers a replicable model for other LMICs to evaluate microbiology systems and align laboratory practices with global AMR control strategies for *Salmonella* infections.

Limitations include;

(i) MDR profiles were inferred from phenotypic data without confirmatory genotypic or plasmid-based analysis.
(ii) Most *Salmonella* isolates were not identified to species or serovar level, restricting lineage-specific interpretations and outbreak mapping.
(iii) Missing patient-level clinical data (e.g., HIV status, antibiotic exposure, comorbidities) limited the ability to link resistance with clinical outcomes.
(iv) Inter-laboratory differences in methods and quality assurance may have influenced *Salmonella* culture positivity and AMR reporting, introducing bias.
(v) Despite identifying One Health gaps, the study did not include data from animal or environmental sources, limiting source attribution.
(vi) Findings may not fully capture the recent national trends in Nigeria, especially in rural or informal health settings not represented in the dataset or recent years.
(vii) Potential underreporting of negative cultures could skew specimen trends for *Salmonella*.

Despite these limitations, our study provides valuable microbiology-focused analysis of *Salmonella* infections diagnostics and resistance trends in a West-African context. Its strength lies in transforming routine laboratory data into actionable insights and revealing systemic microbiological blind spots previously unreported. The translational framework and scalable recommendations make it a timely and impactful contribution to global AMR response efforts in new microbiological perspectives.

## Conclusions

This study successfully identified critical weaknesses in stool specimen collection, species-level identification for *Salmonella* infections, and AST that compromise the detection, reporting, and control of *Salmonella* infections in endemic settings. For *Salmonella* infections, enhancing stool culture prioritization, enforcing species-level identification, and integrating real-time resistance surveillance (e.g., WHONET-linked modules) are urgent and feasible actions. By linking these diagnostic and surveillance failures with escalating resistance to first-line and second-line antibiotics, our study fills a major gap in contextualizing laboratory performance within the broader AMR crisis for *Salmonella* infections. Future work should build on this framework to expand One Health AMR monitoring, incorporate molecular diagnostics, and strengthen laboratory networks to translate microbiological data into early warning and containment systems for *Salmonella* infections and other enteric pathogens.

## Data Availability

All data produced are available online.

https://aslm.org/wp-content/uploads/2023/07/AMR_REPORT_NIGERIA.pdf?x89467

## Acknowledgments

We would like to thank and acknowledge the Mapping Antimicrobial Resistance and Antimicrobial Use Partnership (MAAP) consortium by enhancing global health research by facilitating the public access of all national AMR surveillance reports, including the Nigerian AMR report which was the source of our data collection. We also appreciate the contributions of Nigeria Federal Ministry of Health, the Fleming Fund Country Grant (Phase 1) team, national AMR coordinators, microbiologists, and data managers who ensured quality-assured reporting across surveillance sites in the report.

## References

1. WHO. Interventions for the control of non-typhoidal Salmonella spp. in beef and pork: meeting report and systematic review. Microbiological risk assessment series, 30;. 9789241565240 (WHO) 9789251090497 (FAO)1726-5274. Geneva: 2016. (https://iris.who.int/handle/10665/249529).

2. Marchello CS, Birkhold M, Crump JA, Martin LB, Ansah MO, Breghi G, et al. Complications and mortality of non-typhoidal salmonella invasive disease: a global systematic review and meta-analysis. The Lancet Infectious Diseases 2022;22(5):692–705. DOI: 10.1016/S1473-3099(21)00615-0.

3. Davies MR, Duchene S, Valcanis M, Jenkins AP, Jenney A, Rosa V, et al. Genomic epidemiology of <em>Salmonella</em> Typhi in Central Division, Fiji, 2012 to 2016. The Lancet Regional Health – Western Pacific 2022;24. DOI:10.1016/j.lanwpc.2022.100488.

4. da Silva KE, Tanmoy AM, Pragasam AK, Iqbal J, Sajib MSI, Mutreja A, et al. The international and intercontinental spread and expansion of antimicrobial-resistant <em>Salmonella</em> Typhi: a genomic epidemiology study. The Lancet Microbe 2022;3(8):e567–e577. DOI: 10.1016/S2666-5247(22)00093-3.

5. He Y, Jia Q, Cai K, Xu S, Li H, Xie Q, et al. The global, regional, and national burden of Invasive Non-typhoidal Salmonella (iNTS): An analysis from the Global Burden of Disease Study 1990–2021. PLOS Neglected Tropical Diseases 2025;19(4):e0012960. DOI: 10.1371/journal.pntd.0012960.

6. Stanaway JD, Parisi A, Sarkar K, Blacker BF, Reiner RC, Hay SI, et al. The global burden of non-typhoidal salmonella invasive disease: a systematic analysis for the Global Burden of Disease Study 2017. The Lancet Infectious Diseases 2019;19(12):1312–1324. DOI: 10.1016/S1473-3099(19)30418-9.

7. Thilliez G, Mashe T, Chaibva BV, Robertson V, Bawn M, Tarupiwa A, et al. Population structure of <em>Salmonella enterica</em> Typhi in Harare, Zimbabwe (2012&#x2013;19) before typhoid conjugate vaccine roll-out: a genomic epidemiology study. The Lancet Microbe 2023;4(12):e1005–e1014. DOI: 10.1016/S2666-5247(23)00214-8.

8. Jibril AH, Okeke IN, Dalsgaard A, Olsen JE. Association between antimicrobial usage and resistance in Salmonella from poultry farms in Nigeria. BMC Vet Res 2021;17(1):234. (In eng). DOI: 10.1186/s12917-021-02938-2.

9. Adetunji VO, Davies A, Chisnall T, Ndahi MD, Fagbamila IO, Ekeng E, et al. Genomic Diversity and Antibiotic Resistance of Escherichia coli and Salmonella from Poultry Farms in Oyo State, Nigeria. Microorganisms DOI: 10.3390/microorganisms13061174

10. Batool R, Qamar ZH, Salam RA, Yousafzai MT, Ashorn P, Qamar FN. Efficacy of typhoid vaccines against culture-confirmed S <em>almonella</em> Typhi in typhoid endemic countries: a systematic review and meta-analysis. The Lancet Global Health 2024;12(4):e589–e598. DOI: 10.1016/S2214-109X(23)00606-X.

11. MAAP. ANTIMICROBIAL RESISTANCE SURVEILLANCE GUIDANCE FOR THE AFRICAN REGION. 2024. (https://africacdc.org/download/antimicrobial-resistance-surveillance-guidance-for-the-african-region/).

12. Wang J, Li Q, Jiang Y, Wang Z, Jiao X. <em>fosA7</em>: a silent fosfomycin resistance gene in <em>Salmonella</em>? The Lancet Microbe 2024;5(3):e211. DOI: 10.1016/S2666-5247(23)00342-7.

13. Monte DFM, Doi Y, Lincopan N. High prevalence and global distribution of fosfomycin resistance genes in <em>Salmonella</em> serovars. The Lancet Microbe 2023;4(12):e968. DOI: 10.1016/S2666-5247(23)00261-6.

14. Sati NM, Card RM, Barco L, Muhammad M, Luka PD, Chisnall T, et al. Antimicrobial Resistance and Phylogenetic Relatedness of Salmonella Serovars in Indigenous Poultry and Their Drinking Water Sources in North Central Nigeria. Microorganisms DOI:10.3390/microorganisms12081529.

15. Thilliez G, Kingsley RA. <em>Salmonella</em> intracellular adaptation is key to understand cephalosporin treatment relapse. eBioMedicine 2020;56. DOI: 10.1016/j.ebiom.2020.102802.

16. Alabi ED, Rabiu AG, Adesoji AT. A review of antimicrobial resistance challenges in Nigeria: The need for a one health approach. One Health 2025;20:101053. (In eng). DOI: 10.1016/j.onehlt.2025.101053.

17. Awulu OA, Jenkins A, Balogun BA, Chukwu EE, Fasina FO, Egwuenu A, et al. Prioritising intervention areas for antimicrobial resistance in Nigeria’s human and animal health sectors using a mixed-methods approach. One Health 2025;20:101082. DOI: 10.1016/j.onehlt.2025.101082.

18. Ondoa P, Kapoor G, Alimi Y, Shumba E, Osena G, Maina M, et al. Bacteriology testing and antimicrobial resistance detection capacity of national tiered laboratory networks in sub-Saharan Africa: an analysis from 14&#xa0;countries. The Lancet Microbe 2025;6(1). DOI: 10.1016/j.lanmic.2024.100976.

19. Musa K, Okoliegbe I, Abdalaziz T, Aboushady AT, Stelling J, Gould IM. Laboratory Surveillance, Quality Management, and Its Role in Addressing Antimicrobial Resistance in Africa: A Narrative Review. Antibiotics (Basel) 2023;12(8) (In eng). DOI: 10.3390/antibiotics12081313.

20. Bertagnolio S, Dobreva Z, Centner CM, Olaru ID, Donà D, Burzo S, et al. WHO global research priorities for antimicrobial resistance in human health. The Lancet Microbe 2024;5(11). DOI: 10.1016/S2666-5247(24)00134-4.

21. Wang Y, Xu X, Jia S, Qu M, Pei Y, Qiu S, et al. A global atlas and drivers of antimicrobial resistance in Salmonella during 1900-2023. Nature communications 2025;16(1):4611. DOI: 10.1038/s41467-025-59758-3.

22. Mao S, Soputhy C, Lay S, Jacobs J, Ku GM, Chau D, et al. The barriers and facilitators of implementing a national laboratory-based AMR surveillance system in Cambodia: key informants’ perspectives and assessments of microbiology laboratories. Front Public Health 2023;11:1332423. (In eng). DOI: 10.3389/fpubh.2023.1332423.

23. Ashton PM, Chunga Chirambo A, Meiring JE, Patel PD, Mbewe M, Silungwe N, et al. Evaluating the relationship between ciprofloxacin prescription and non-susceptibility in <em>Salmonella</em> Typhi in Blantyre, Malawi: an observational study. The Lancet Microbe 2024;5(3):e226–e234. DOI: 10.1016/S2666-5247(23)00327-0.

24. Castanheira S, López-Escarpa D, Pucciarelli MG, Cestero JJ, Baquero F, García-del Portillo F. An alternative penicillin-binding protein involved in <em>Salmonella</em> relapses following ceftriaxone therapy. eBioMedicine 2020;55. DOI: 10.1016/j.ebiom.2020.102771.

25. Zhou l, Wang M, Li W. Genomic epidemiology of <em>Salmonella</em>: the need to consider vaccination history and nutritional status in resistance transmission studies. The Lancet Microbe 2025;6(2). DOI: 10.1016/j.lanmic.2024.101011.

26. Browne AJ, Chipeta MG, Fell FJ, Haines-Woodhouse G, Kashef Hamadani BH, Kumaran EAP, et al. Estimating the subnational prevalence of antimicrobial resistant <em>Salmonella enterica</em> serovars Typhi and Paratyphi A infections in 75 endemic countries, 1990&#x2013;2019: a modelling study. The Lancet Global Health 2024;12(3):e406–e418. DOI: 10.1016/S2214-109X(23)00585-5.

27. Piña-Iturbe A, Díaz-Gavidia C, Álvarez FP, Barron-Montenegro R, Álvarez-Espejo DM, García P, et al. Genomic characterisation of the population structure and antibiotic resistance of <em>Salmonella enterica</em> serovar Infantis in Chile, 2009&#x2013;2022. The Lancet Regional Health – Americas 2024;32. DOI: 10.1016/j.lana.2024.100711.

28. Schatten H. Salmonella Methods and Protocols. Hatfield, Hertfordshire, UK: Springer Nature, 2021. 10.1007/978-1-0716-0791-6.

29. Huang C. Salmonella - Current Trends and Perspectives in Detection and Control2024. DOI: 10.5772/intechopen.1001722

30. Chiou CS, Hong YP, Wang YW, Chen BH, Teng RH, Song HY, et al. Antimicrobial Resistance and Mechanisms of Azithromycin Resistance in Nontyphoidal Salmonella Isolates in Taiwan, 2017 to 2018. Microbiol Spectr 2023;11(1):e0336422. (In eng). DOI: 10.1128/spectrum.03364-22.

31. Peruzy MF, Murru N, Carullo MR, La Tela I, Rippa A, Balestrieri A, et al. Antibiotic-Resistant Salmonella Circulation in the Human Population in Campania Region (2010–2023). Antibiotics DOI:10.3390/antibiotics14020189.

32. Talukder H, Roky SA, Debnath K, Sharma B, Ahmed J, Roy S. Prevalence and Antimicrobial Resistance Profile of Salmonella Isolated from Human, Animal and Environment Samples in South Asia: A 10-Year Meta-analysis. Journal of Epidemiology and Global Health 2023;13(4):637–652. DOI: 10.1007/s44197-023-00160-x.

33. Dyson ZA, Ashton PM, Khanam F, Chunga Chirambo A, Shakya M, Meiring JE, et al. Pathogen diversity and antimicrobial resistance transmission of <em>Salmonella enterica</em> serovars Typhi and Paratyphi A&#xa0;in Bangladesh, Nepal, and Malawi: a genomic epidemiological study. The Lancet Microbe 2024;5(8). DOI: 10.1016/S2666-5247(24)00047-8.

34. Hu Y, Wang J, Chen S, Li Y, Zhang R, Chen S. Non-typhoidal salmonella invasive infections in China. The Lancet Infectious Diseases 2022;22(7):939. DOI: 10.1016/S1473-3099(22)00347-4.

35. Amuasi JH, May J. Non-typhoidal salmonella: invasive, lethal, and on the loose. The Lancet Infectious Diseases 2019;19(12):1267–1269. DOI: 10.1016/S1473-3099(19)30521-3.

36. Medugu N, Michelow IC, Poole C, Obaro SK. Azithromycin mass drug administration: balancing survival benefits and risks in children. The Lancet Infectious Diseases. DOI: 10.1016/S1473-3099(25)00363-9.

37. Aleksandrowicz A, Carolak E, Dutkiewicz A, Błachut A, Waszczuk W, Grzymajlo K. Better together-Salmonella biofilm-associated antibiotic resistance. Gut Microbes 2023;15(1):2229937. (In eng). DOI: 10.1080/19490976.2023.2229937.

38. Griewisch KF, Pierce JG, Elfenbein JR. Genetic Determinants of Salmonella Resistance to the Biofilm-Inhibitory Effects of a Synthetic 4-Oxazolidinone Analog. Appl Environ Microbiol 2020;86(20) (In eng). DOI: 10.1128/aem.01120-20.

39. Osena G, Kapoor G, Kalanxhi E, Ouassa T, Shumba E, Brar S, et al. Antimicrobial resistance in Africa: A retrospective analysis of data from 14 countries, 2016–2019. PLOS Medicine 2025;22(6):e1004638. DOI: 10.1371/journal.pmed.1004638.

40. Skrivankova VW, Richmond RC, Woolf BAR, Yarmolinsky J, Davies NM, Swanson SA, et al. Strengthening the Reporting of Observational Studies in Epidemiology Using Mendelian Randomization: The STROBE-MR Statement. JAMA: the journal of the American Medical Association 2021;326(16):1614–1621. (In eng). DOI:10.1001/jama.2021.18236.

41. Kajumbula HM, Amoako DG, Tessema SK, Aworh MK, Chikuse F, Okeke IN, et al. Enhancing clinical microbiology for genomic surveillance of antimicrobial resistance implementation in Africa. Antimicrobial Resistance & Infection Control 2024;13(1):135. DOI: 10.1186/s13756-024-01472-8.

42. WHO. Global Antimicrobial Resistance and Use Surveillance System (GLASS) Report. 2021. (https://iris.who.int/bitstream/handle/10665/341666/9789240027336-eng.pdf?sequence=1).

43. Igbinosa EO, Beshiru A, Igbinosa IH, Okoh AI. Antimicrobial resistance and genetic characterisation of Salmonella enterica from retail poultry meats in Benin City, Nigeria. LWT 2022;169:114049. DOI: 10.1016/j.lwt.2022.114049.

44. Ahmed AO, Raji MA, Mamman PH, Kwanashie CN, Raufu IA, Aremu A, et al. Salmonellosis: Serotypes, prevalence and multi-drug resistant profiles of Salmonella enterica in selected poultry farms, Kwara State, North Central Nigeria. Onderstepoort J Vet Res 2019;86(1):e1–e8. (In eng). DOI: 10.4102/ojvr.v86i1.1667.

45. Dlamini SB, Mlambo V, Mnisi CM, Ateba CN. Virulence, multiple drug resistance, and biofilm-formation in Salmonella species isolated from layer, broiler, and dual-purpose indigenous chickens. PloS one 2024;19(10):e0310010. (In eng). DOI:10.1371/journal.pone.0310010.

46. Akinyemi KO, Bamiro BS, Coker AO. Salmonellosis in Lagos, Nigeria: incidence of Plasmodium falciparum-associated co-infection, patterns of antimicrobial resistance, and emergence of reduced susceptibility to fluoroquinolones. J Health Popul Nutr 2007;25(3):351–8. (In eng). PMID: 18330069. PMCID: PMC2754035.

47. Lu X, Zeng M, Xu J, Zhou H, Gu B, Li Z, et al. Epidemiologic and genomic insights on <em>mcr-1</em>-harbouring <em>Salmonella</em> from diarrhoeal outpatients in Shanghai, China, 2006&#x2013;2016. eBioMedicine 2019;42:133–144. DOI:10.1016/j.ebiom.2019.03.006.

48. Pijnacker R, Dallman TJ, Tijsma ASL, Hawkins G, Larkin L, Kotila SM, et al. An international outbreak of <em>Salmonella enterica</em> serotype Enteritidis linked to eggs from Poland: a microbiological and epidemiological study. The Lancet Infectious Diseases 2019;19(7):778–786. DOI: 10.1016/S1473-3099(19)30047-7.

49. Qamar FN, Yousafzai MT, Khalid M, Kazi AM, Lohana H, Karim S, et al. Outbreak investigation of ceftriaxone-resistant <em>Salmonella enterica</em> serotype Typhi and its risk factors among the general population in Hyderabad, Pakistan: a matched case-control study. The Lancet Infectious Diseases 2018;18(12):1368–1376. DOI: 10.1016/S1473-3099(18)30483-3.

50. Olufon O, Seale AC, Iyanger N, Wynne-Evans E. Use of whole-genome sequencing for public health intervention: outbreak investigation of a cluster of cases of salmonella foodborne illness in England, 2016. The Lancet 2018;392:S10. DOI:10.1016/S0140-6736(18)32081-6.

51. Zheng B, Feng Y. MCR-1-producing <em>Salmonella</em> Typhimurium ST34 links animal foods to human community infections. eBioMedicine 2019;42:10–11. DOI: 10.1016/j.ebiom.2019.03.073.

52. Marks F, Im J, Park SE, Pak GD, Jeon HJ, Wandji Nana LR, et al. Incidence of typhoid fever in Burkina Faso, Democratic Republic of the Congo, Ethiopia, Ghana, Madagascar, and Nigeria (the Severe Typhoid in Africa programme): a population-based study. The Lancet Global Health 2024;12(4):e599–e610. DOI: 10.1016/S2214-109X(24)00007-X.

53. Kariuki S, Onsare RS. High burden of typhoid disease in sub-Saharan Africa calls for urgent roll-out of typhoid conjugate vaccines. The Lancet Global Health 2024;12(4):e537–e538. DOI: 10.1016/S2214-109X(24)00079-2.

54. Sanni AO, Onyango J, Rota AF, Mikecz O, Usman A, PicaCiamarra U, et al. Underestimated economic and social burdens of non-Typhoidal Salmonella infections: The One Health perspective from Nigeria. One Health 2023;16:100546. DOI: 10.1016/j.onehlt.2023.100546.

55. Sanni AO, Jibril AH, Fasanmi OG, Adebowale OO, Jambalang AR, Shittu A, et al. Non-typhoidal Salmonella in Nigeria: do outcomes of ‘multisectoral’ surveillance, treatment and control justify the intervention costs? International Journal of Veterinary Science and Medicine 2024;12(1):48–59. DOI: 10.1080/23144599.2024.2365567.

56. Dagah H, Ameh JA, Mailafia S, Dantong DD, Onigbanjo OH, Ifeanye CIC, et al. Prevalence and antimicrobial susceptibility of Salmonella from roasted meat ("Suya") sold in federal capital territory, Abuja, Nigeria. The Microbe 2024;5:100179. DOI: 10.1016/j.microb.2024.100179.

57. Akinyemi KO, Fakorede CO, Linde J, Methner U, Wareth G, Tomaso H, et al. Whole genome sequencing of Salmonella enterica serovars isolated from humans, animals, and the environment in Lagos, Nigeria. BMC Microbiology 2023;23(1):164. DOI: 10.1186/s12866-023-02901-1.

58. Aworh MK, Nilsson P, Egyir B, Owusu FA, Hendriksen RS. Rare serovars of non-typhoidal Salmonella enterica isolated from humans, beef cattle and abattoir environments in Nigeria. PloS one 2024;19(1):e0296971. DOI:10.1371/journal.pone.0296971.

59. Ikhimiukor OO, Oaikhena AO, Afolayan AO, Fadeyi A, Kehinde A, Ogunleye VO, et al. Genomic characterization of invasive typhoidal and non-typhoidal Salmonella in southwestern Nigeria. PLOS Neglected Tropical Diseases 2022;16(8):e0010716. DOI: 10.1371/journal.pntd.0010716.

60. Igbinosa IH, Amolo CN, Beshiru A, Akinnibosun O, Ogofure AG, El-Ashker M, et al. Identification and characterization of MDR virulent Salmonella spp isolated from smallholder poultry production environment in Edo and Delta States, Nigeria. PloS one 2023;18(2):e0281329. DOI: 10.1371/journal.pone.0281329.

61. Okeke IN, de Kraker MEA, Van Boeckel TP, Kumar CK, Schmitt H, Gales AC, et al. The scope of the antimicrobial resistance challenge. The Lancet 2024;403(10442):2426–2438. DOI: 10.1016/S0140-6736(24)00876-6.

62. Kyu HH, Vongpradith A, Dominguez R-MV, Ma J, Albertson SB, Novotney A, et al. Global, regional, and national age-sex-specific burden of diarrhoeal diseases, their risk factors, and aetiologies, 1990&#x2013;2021, for 204 countries and territories: a systematic analysis for the Global Burden of Disease Study 2021. The Lancet Infectious Diseases 2025;25(5):519–536. DOI: 10.1016/S1473-3099(24)00691-1.

63. Naghavi M, Vollset SE, Ikuta KS, Swetschinski LR, Gray AP, Wool EE, et al. Global burden of bacterial antimicrobial resistance 1990&#x2013;2021: a systematic analysis with forecasts to 2050. The Lancet 2024;404(10459):1199–1226. DOI:10.1016/S0140-6736(24)01867-1.

64. Mkangara M. Prevention and Control of Human Salmonella enterica Infections: An Implication in Food Safety. International Journal of Food Science 2023;2023(1):8899596. DOI: 10.1155/2023/8899596.

65. McConn BR, Kraft AL, Durso LM, Ibekwe AM, Frye JG, Wells JE, et al. An analysis of culture-based methods used for the detection and isolation of Salmonella spp., Escherichia coli, and Enterococcus spp. from surface water: A systematic review. Science of The Total Environment 2024;927:172190. 10.1016/j.scitotenv.2024.172190.

66. Marami D, Hailu K, Tolera M. Prevalence and antimicrobial susceptibility pattern of Salmonella and Shigella species among asymptomatic food handlers working in Haramaya University cafeterias, Eastern Ethiopia. BMC research notes 2018;11(1):74. DOI: 10.1186/s13104-018-3189-9.

67. Dade Y, Kannaiyan M, Dedecha W, Daka D, Husen O, Gemechu T, et al. Prevalence, antimicrobial susceptibility pattern and associated factors of Salmonella and Shigella among under five children with diarrhea attending Bule Hora University Teaching Hospital, Bule Hora, West Guji, Ethiopia. BMC infectious diseases 2025;25(1):571. (In eng). DOI: 10.1186/s12879-025-10960-0.

68. Matee M, Mshana SE, Mtebe M, Komba EV, Moremi N, Lutamwa J, et al. Mapping and gap analysis on antimicrobial resistance surveillance systems in Kenya, Tanzania, Uganda and Zambia. Bulletin of the National Research Centre 2023;47(1):12. DOI:10.1186/s42269-023-00986-2.

69. Kanzi AM, Smith SI, Msefula C, Mwaba J, Ajayi A, Kwenda G, et al. Expediting pathogen genomics adoption for enhanced foodborne disease surveillance in Africa. eBioMedicine 2025;111. DOI: 10.1016/j.ebiom.2024.105500.

70. Ahmad R, Zhu N, Jain R, Joshi J, Mpundu M, Gutierrez P, et al. Systems Policy Analysis for Antimicrobial Resistance Targeted Action (SPAARTA): A Research Protocol [version 1; peer review: 1 approved, 1 approved with reservations]. Wellcome Open Research 2024;9(700). DOI: 10.12688/wellcomeopenres.22923.1.

